# Thyroid Autoimmunity Does Not Delineate a Cardiometabolic or Androgenic Phenotype in Women With Polycystic Ovary Syndrome: A Pre-Specified Cross-Sectional Analysis

**DOI:** 10.64898/2026.03.17.26348579

**Authors:** Natalia Piórkowska, Lech Madeyski, Marcin Leśniewski, Grzegorz Franik, Anna Bizoń

**Author notes:** Corresponding author: Natalia Piórkowska, PhD.

## Abstract

**Background:** Thyroid autoimmunity (TAI) is frequently reported in women with polycystic ovary syndrome (PCOS), yet its clinical relevance for cardiometabolic and androgenic severity remains uncertain. We evaluated whether TAI identifies a metabolically or androgenically more severe PCOS phenotype using pre-specified exposure definitions and cardiometabolic endpoints.

**Methods:** This cross-sectional study included 1,300 women with confirmed PCOS in the source dataset. Thyroid autoimmunity was defined a priori using three definitions: anti-thyroid peroxidase antibodies above the laboratory upper limit of normal (TAI_A, primary definition), anti-TPO positivity combined with thyroid-stimulating hormone >4.0 mIU/L (TAI_B), and high-titer anti-TPO >100 IU/mL (TAI_C). The primary endpoint was triglyceride-to-high-density lipoprotein cholesterol ratio (TG/HDL-C) >3.5. Secondary endpoints included non-HDL-C ≥130 mg/dL and 120-minute oral glucose tolerance test (OGTT) glucose ≥140 mg/dL. Associations were assessed using age-adjusted Firth logistic regression models in complete-case cohorts. Sensitivity analyses included restriction to euthyroid participants, alternative TAI definitions, trimming of extreme values (1–99%), and bootstrap-based confidence intervals. Exploratory hormonal comparisons were adjusted using the Benjamini–Hochberg false discovery rate.

**Results:** TAI_A was not significantly associated with the primary endpoint (TG/HDL >3.5) (OR 0.77, 95% CI 0.21–1.67). No significant associations were observed for secondary endpoints including non-HDL-C ≥130 mg/dL (OR 1.09, 95% CI 0.61–1.76) or impaired glucose tolerance on OGTT (OR 1.27, 95% CI 0.63–2.18). Results remained directionally consistent across alternative TAI definitions and sensitivity analyses, including restriction to euthyroid women and trimming of extreme values. In exploratory analyses, thyroid-stimulating hormone levels differed between TAI-positive and TAI-negative women, while no androgenic or cardiometabolic parameters remained significant after false discovery rate correction. Model diagnostics did not indicate major violations of model assumptions.

**Conclusion:** In this large cross-sectional cohort of women with PCOS, thyroid autoimmunity was not associated with an adverse cardiometabolic or androgenic phenotype. Anti-TPO positivity alone therefore does not appear to identify a metabolically high-risk PCOS subgroup under the studied conditions. Prospective studies are needed to clarify the longitudinal implications of thyroid autoimmunity in PCOS.

## 3. Introduction

Polycystic ovary syndrome (PCOS) is the most common endocrine disorder in women of reproductive age and is characterized by hyperandrogenism, ovulatory dysfunction, and polycystic ovarian morphology (1). Beyond reproductive features, PCOS is increasingly recognized as a heterogeneous metabolic condition associated with insulin resistance, dyslipidemia, and increased cardiometabolic risk (2). Considerable effort has therefore been directed toward identifying clinically meaningful PCOS subphenotypes that may differ in metabolic severity and long-term risk profiles (3,4).

Thyroid autoimmunity (TAI), most commonly defined by the presence of anti–thyroid peroxidase antibodies (anti-TPO), has been reported to occur more frequently in women with PCOS compared with the general population (5,6). Autoimmune thyroid disease has been reported in 18–40% of PCOS women, depending on PCOS diagnostic criteria and ethnicity (6). This co-occurrence has raised interest in potential shared mechanisms, including chronic low-grade inflammation, immune dysregulation, and interactions between metabolic and endocrine pathways (7–9). The concept of an immune–metabolic axis has gained traction in recent years, suggesting that autoimmune activity could contribute to metabolic dysfunction through inflammatory signaling, alterations in insulin sensitivity, or modulation of steroidogenesis. Within this framework, it has been hypothesized that thyroid autoimmunity might identify a subset of women with PCOS characterized by greater metabolic or androgenic severity.

However, the clinical implications of thyroid autoimmunity in PCOS remain unclear. Previous studies have produced inconsistent findings, with some reports suggesting associations between TAI and adverse lipid profiles, insulin resistance, or impaired glucose tolerance, while others have failed to demonstrate such relationships (9,10). Many of these studies were limited by small sample sizes, heterogeneous definitions of thyroid autoimmunity, varying thyroid functional states, or insufficient adjustment strategies. Furthermore, few investigations have evaluated cardiometabolic endpoints using clearly pre-specified thresholds or conducted robustness analyses across alternative definitions of TAI. As a result, whether thyroid autoimmunity meaningfully delineates a metabolically or androgenically more severe PCOS phenotype remains unresolved.

The present study aimed to address this gap using a pre-specified cross-sectional design in a large cohort of women with PCOS. Thyroid autoimmunity was defined a priori using three operational definitions, including anti-TPO positivity above the laboratory upper limit of normal and a high-titer threshold. The primary endpoint was an elevated triglyceride-to-high-density lipoprotein cholesterol ratio (TG/HDL-C >3.5), selected as a clinically relevant marker of atherogenic risk and insulin resistance. Secondary endpoints included non–HDL-C ≥130 mg/dL and impaired glucose tolerance defined by 120-minute oral glucose tolerance test (OGTT) glucose ≥140 mg/dL. We hypothesized that thyroid autoimmunity would be associated with a more adverse cardiometabolic and androgenic profile. To ensure robustness, analyses incorporated predefined sensitivity strategies and model diagnostics.

## 4. Materials and Methods

### 4.1 Study Design and Population

This study was a pre-specified cross-sectional analysis of women with PCOS evaluated at a tertiary endocrine referral center. The analysis used an existing clinical dataset comprising women who underwent standardized clinical, biochemical, and metabolic assessment as part of routine care.

PCOS was diagnosed according to the Rotterdam criteria, requiring the presence of at least two of the following features after exclusion of other etiologies: (1) oligo- or anovulation, (2) clinical or biochemical hyperandrogenism, and (3) polycystic ovarian morphology on ultrasound.

The source dataset included 1,300 women with confirmed PCOS. Participants were eligible for the present analysis if measurements of anti-TPO were available to define thyroid autoimmunity status. Women with missing anti-TPO measurements were excluded from analyses requiring TAI classification.

For each endpoint analysis, complete-case cohorts were defined based on the availability of variables required to construct the corresponding outcome and covariates. Consequently, the number of analyzed participants varied across endpoints. The selection process from the source dataset to endpoint-specific analytic cohorts is illustrated in the STROBE participant flow diagram.

The study was conducted in accordance with the principles of the Declaration of Helsinki. Ethical approval was obtained from the institutional review board of the participating center (approval number: to be inserted). As this study involved secondary analysis of routinely collected clinical data, informed consent procedures followed local regulatory requirements.

### 4.2 Clinical and Laboratory Assessment

All participants underwent standardized clinical evaluation and fasting venous blood sampling as part of routine clinical assessment at the study center.

Laboratory measurements included the following domains:

#### Thyroid parameters

Thyroid-stimulating hormone (TSH), free thyroxine (FT4), and anti-TPO.

#### Lipid profile

Total cholesterol (TC), high-density lipoprotein cholesterol (HDL-C), low-density lipoprotein cholesterol (LDL-C), and triglycerides (TG).

Derived lipid indices were calculated as follows: non–HDL cholesterol (non–HDL-C) was defined as TC minus HDL-C, and the triglyceride-to-HDL cholesterol ratio (TG/HDL-C) was calculated as TG divided by HDL-C.

#### Glucose metabolism

Fasting plasma glucose and 120-minute plasma glucose measured during a standard 75-g oral glucose tolerance test (OGTT).

#### Androgen-related parameters

Total testosterone (TT), free testosterone (FT), dehydroepiandrosterone sulfate (DHEAS), and other routinely measured reproductive hormones when available.

All biochemical analyses were performed in the same certified clinical laboratory using standardized assays implemented in routine clinical practice.

### 4.3 Definition of Thyroid Autoimmunity

Thyroid autoimmunity (TAI) was defined a priori using three pre-specified definitions based on anti-TPO and TSH levels.

#### Primary definition (TAI_A)

The primary exposure definition (TAI_A) was defined as anti-TPO concentration above the upper limit of normal (ULN) according to the reference range of the local laboratory performing the assays.

#### Alternative definitions

Two additional definitions of thyroid autoimmunity were evaluated in sensitivity analyses:

● TAI_B: anti-TPO above ULN combined with elevated TSH (>4.0 mIU/L), representing autoimmune thyroid disease with biochemical thyroid dysfunction.
● TAI_C: high-titer anti-TPO positivity (anti-TPO >100 IU/mL), representing a more stringent definition of thyroid autoimmunity.

All primary analyses used TAI_A as the main exposure variable, whereas TAI_B and TAI_C were examined in sensitivity analyses to evaluate the robustness of associations across alternative definitions of thyroid autoimmunity.

### 4.4 Cardiometabolic and Androgenic Endpoints Primary endpoint

The pre-specified primary endpoint was an elevated TG/HDL-C, defined as TG/HDL-C >3.5. This threshold has been widely used as a marker of atherogenic dyslipidemia and insulin resistance in cardiometabolic risk studies.

#### Secondary endpoints

Secondary cardiometabolic endpoints included:

● non–HDL-C ≥130 mg/dL, and
● 120-minute plasma glucose ≥140 mg/dL during the OGTT.

All cardiometabolic endpoints were analyzed as binary outcomes in regression models.

#### Androgenic parameters

Androgen-related parameters (TT, FT, and DHEAS) were analyzed as continuous variables in exploratory comparisons between TAI-positive and TAI-negative participants. To account for multiple testing, p-values from these exploratory analyses were adjusted using the Benjamini–Hochberg false discovery rate (FDR) procedure.

### 4.5 Data Processing and Quality Control

All data processing procedures were pre-specified and implemented using reproducible analysis scripts.

#### Units and harmonization

Laboratory values were reviewed for consistency of measurement units. When necessary, unit harmonization procedures were applied to ensure comparability across measurements.

#### Parsing and numeric cleaning

Laboratory entries recorded as ranges (e.g., “a–b”) were converted to numeric values using predefined parsing rules. Entries reported as threshold values (e.g., “<x” or “>x”) were conservatively mapped to the corresponding boundary value to preserve numerical interpretability.

#### Derived variables

The cardiometabolic indices TG/HDL-C and non–HDL-C were calculated as described above.

#### Transformation and trimming

Variables with right-skewed distributions (e.g., anti-TPO or TG) were evaluated using log-transformation for descriptive and visualization purposes. In sensitivity analyses, extreme values were trimmed at the 1st and 99th percentiles (1–99% trimming) to assess the robustness of regression estimates.

#### Missing data

Patterns of missingness were evaluated descriptively. Primary analyses were conducted using complete-case data for each endpoint-specific model, and the number of analyzed observations therefore varied depending on variable availability. No imputation procedures were performed.

#### Outlier and influence assessment

Potentially influential observations were evaluated using standardized residuals and Cook’s distance diagnostics within regression models. These diagnostics were used to assess model stability rather than to automatically exclude observations.

### 4.6 Statistical Analysis

All statistical analyses were performed using Python (version 3.12) with standard scientific libraries (NumPy, pandas, SciPy, statsmodels, and scikit-learn). All tests were two-sided, and p-values <0.05 were considered statistically significant unless otherwise specified.

#### 4.6.1 Primary analysis

The association between thyroid autoimmunity (TAI_A) and the primary endpoint (TG/HDL-C >3.5) was assessed using Firth logistic regression, which reduces small-sample bias in logistic models.

Models were adjusted for age as a minimal a priori confounder. Effect estimates are reported as odds ratios (ORs) with 95% confidence intervals (CIs). Confidence intervals were obtained using bootstrap resampling to provide robust interval estimation.

#### 4.6.2 Secondary analyses

Separate regression models were fitted for the secondary cardiometabolic endpoints (non–HDL-C ≥130 mg/dL and OGTT 120-minute glucose ≥140 mg/dL), using the same modeling strategy and age adjustment as in the primary analysis.

Selected metabolic and hormonal parameters were additionally evaluated in exploratory continuous analyses comparing TAI-positive and TAI-negative groups.

#### 4.6.3 Sensitivity analyses

Robustness of the primary findings was assessed using multiple pre-specified sensitivity analyses:

● Restriction to euthyroid participants (TSH within the reference range and normal FT4).
● Use of alternative TAI definitions (TAI_B and TAI_C).
● Trimming of extreme values (1st–99th percentiles).
● Bootstrap-based confidence interval estimation.

#### 4.6.4 Multiple testing control

For exploratory comparisons involving multiple hormonal and metabolic parameters, p-values were adjusted using the Benjamini–Hochberg false discovery rate (FDR) procedure.

#### 4.6.5 Model diagnostics

Model calibration was evaluated using calibration plots and the Hosmer–Lemeshow goodness-of-fit test.

Potentially influential observations were examined using Cook’s distance diagnostics. Multicollinearity among predictors was assessed using variance inflation factors (VIFs).

The assumption of linearity for the age covariate was evaluated using spline-based sensitivity models.

Because some standard diagnostic procedures are not directly available for Firth logistic regression, selected diagnostic measures were computed using auxiliary standard logistic models fitted to the same complete-case cohorts.

## 5 Results

### 5.1 Study population characteristics

A total of 1,300 women with confirmed PCOS were included in the source dataset. Measurements of anti-TPO were available for 1,055 participants, allowing classification of thyroid autoimmunity status. Among these, 84 women (8.0%) were classified as TAI-positive according to the primary definition (anti-TPO above the laboratory upper limit of normal), whereas 971 women (92.0%) were classified as TAI-negative.

Baseline clinical and biochemical characteristics stratified by TAI status are summarized in Table 1. Women with TAI showed higher TSH concentrations, consistent with the known physiological relationship between thyroid autoimmunity and thyroid function.

**Table 1.** Baseline characteristics.

| Variable | TAI-positive<br>(n=84) | TAI-negative<br>(n=971) | Missing, n | SMD | P value |
| --- | --- | --- | --- | --- | --- |
| Clinical characteristics |  |  |  |  |  |
| Age (years) | 22 [21, 23] | 22 [20, 23] | 0 | 0.103 | 1.000 |
| Thyroid parameters |  |  |  |  |  |
| TSH (mIU/L) | 2.4 [1.46, 3.52] | 1.77 [1.27, 2.42] | 1 | 0.896 | <0.001 |
| Anti-TPO (IU/mL) | 129 [70.9, 261] | 9.9 [9, 13] | 0 | 4.247 | <0.001 |
| Lipid profile |  |  |  |  |  |
| TG (mg/dL) | 75.3 [58.8, 103] | 77 [58.9, 105] | 2 | -0.018 | 0.718 |
| HDL-C (mg/dL) | 56.6 [48, 66] | 57.8 [48.6, 67.2] | 2 | -0.012 | 0.497 |
| TC (mg/dL) | 164 [137, 188] | 164 [146, 185] | 2 | -0.048 | 0.948 |
| LDL-C (mg/dL) | 85.5 [68.7, 107] | 87.9 [72, 106] | 2 | -0.043 | 0.560 |
| Derived lipid indices |  |  |  |  |  |
| TG/HDL-C ratio | 1.24 [0.95, 2.22] | 1.29 [0.904, 2] | 2 | 0.048 | 0.669 |
| Non-HDL-C (mg/dL) | 103 [81.9, 127] | 104 [86.8, 125] | 2 | -0.043 | 0.677 |
| Glucose metabolism |  |  |  |  |  |
| Glu0 (mg/dL) | 84 [79.9, 89.4] | 83.7 [80.3, 87.9] | 19 | 0.075 | 0.831 |
| OGlu120 (mg/dL) | 106 [93.2, 126] | 108 [91.2, 127] | 20 | 0.031 | 0.693 |
| Androgen-related parameters |  |  |  |  |  |
| TT (ng/mL) | 0.369 [0.258, 0.483] | 0.37 [0.272, 0.517] | 1 | -0.128 | 0.957 |
| FT (pg/mL) | 2.71 [1.41, 3.94] | 2.14 [1.19, 3.65] | 118 | 0.074 | 0.027 |
| DHEAS (µg/mL) | 316 [245, 419] | 320 [245, 413] | 1 | -0.094 | 0.814 |
| Androstenedione (ng/mL) | 1.42 [1.12, 1.76] | 1.48 [1.12, 1.92] | 364 | -0.216 | 0.662 |
| Reproductive hormones |  |  |  |  |  |
| AMH (ng/mL) | 4.88 [4.69, 5.37] | 6.01 [4.53, 10.3] | 990 | -0.516 | 0.516 |
| LH (IU/L) | 6.95 [4.56, 10.5] | 7.21 [4.96, 10.4] | 1 | -0.002 | 0.655 |
| FSH (IU/L) | 5.85 [5.09, 6.73] | 5.86 [5.01, 6.91] | 1 | 0.086 | 0.951 |
| Binary endpoints |  |  |  |  |  |
| TG/HDL-C > 3.5, n (%) | 4/84 (4.8%) | 66/969 (6.8%) | 2 | -0.087 |  |
| Non-HDL-C ≥ 130 (mg/dL), n (%) | 19/84 (22.6%) | 205/969 (21.2%) | 2 | 0.036 |  |
| OGTT 120-min glucose ≥ 140 (mg/dL), n (%) | 14/79 (17.7%) | 142/956 (14.9%) | 20 | 0.056 |  |
| Euthyroid by TSH, n (%) | 69/84 (82.1%) | 937/970 (96.6%) | 1 | -0.465 |  |

Legend: TSH - thyroid-stimulating hormone, Anti\_TPO - anti-thyroid peroxidase antibodies, TG - triglycerides, HDL-C - high-density lipoprotein cholesterol, TC - total cholesterol, LDL-C - low-density lipoprotein cholesterol, glu0 - fasting glucose, glu120 - glucose concentration 120 minutes after oral glucose load, TT- total testosterone, FT - free testosterone, DHEAS - dehydroepiandrosterone sulfate, AMH- Anti-Müllerian hormone, LH - luteinizing hormone, FSH - follicle-stimulating hormone, SHBG - sex hormone-binding globulin.

Distributions of selected thyroid and cardiometabolic variables by TAI status are illustrated in Figure 1.

**Figure 1.**
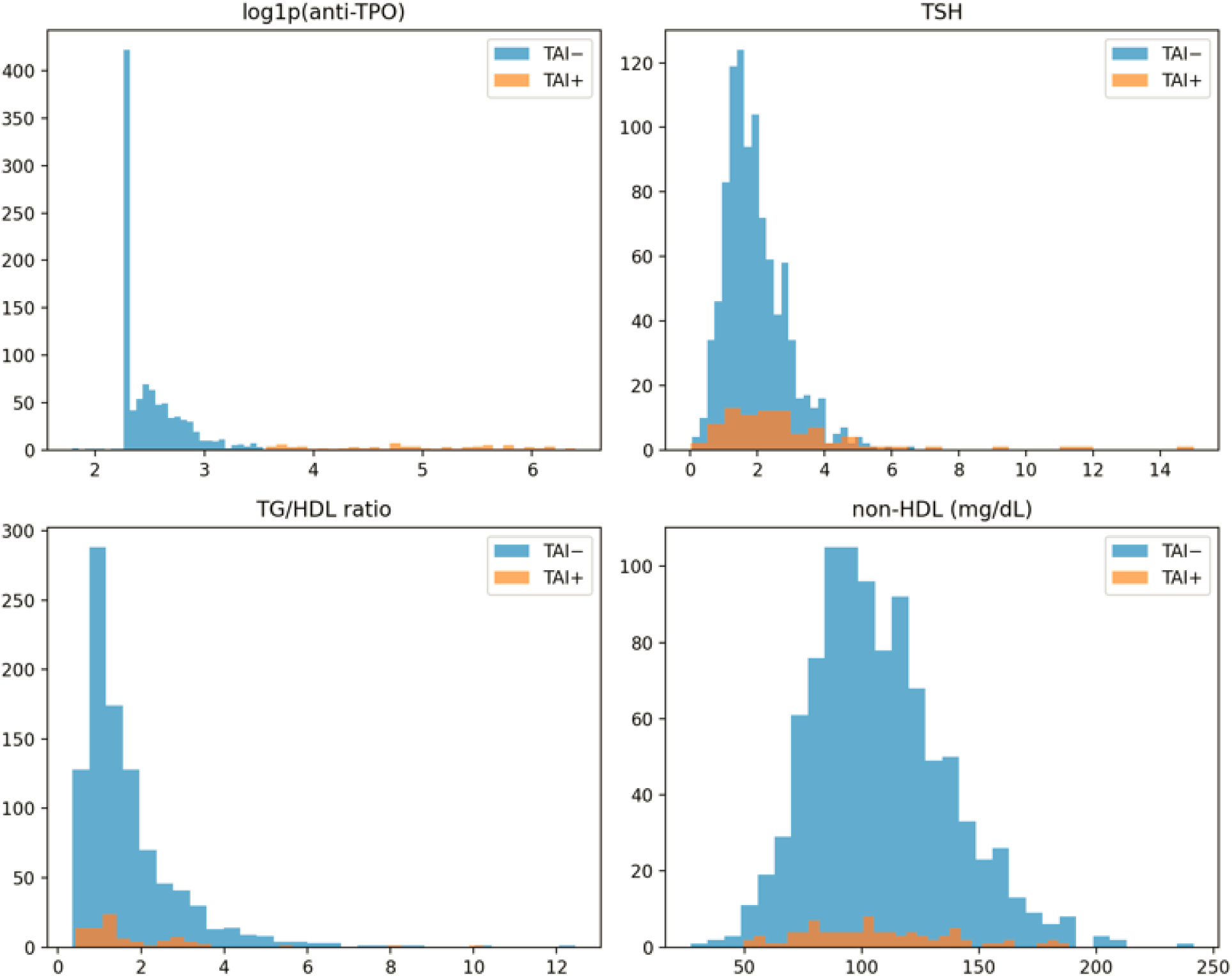
Distribution of thyroid and cardiometabolic.

In contrast, no clinically meaningful differences were observed between TAI-positive and TAI-negative participants for lipid parameters, glucose metabolism markers, or androgen-related variables, including TG, HDL-C, TG/HDL-C ratio, non-HDL-C, fasting glucose, or androgen indices (Figure 2). Exploratory comparisons of multiple biochemical and hormonal markers confirmed that TSH was the only variable remaining statistically significant after false discovery rate correction, whereas no androgenic or cardiometabolic parameters showed significant group differences.

**Figure 2.**
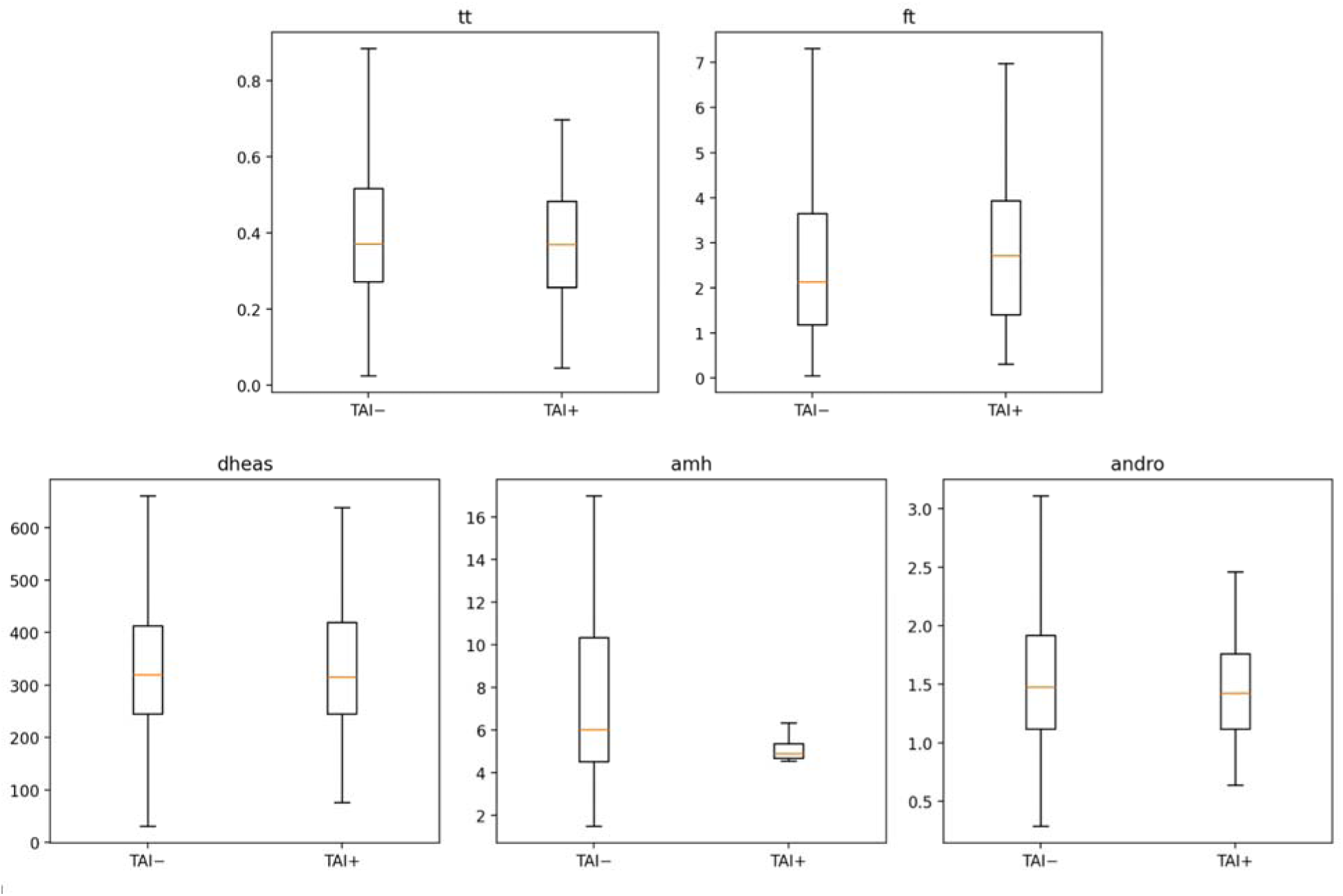
Androgen distributions.

Patterns of missing data were evaluated descriptively. Anti-TPO measurements were available in 1,055 of 1,300 women (81.2%), while other cardiometabolic variables generally had substantially lower missingness. For example, TG, total cholesterol, HDL-C, TSH, and DHEAS had missingness rates below approximately 10%, whereas higher missingness was observed for selected reproductive hormones such as AMH and SHBG. The distribution of missing values across variables is shown in Figure 3.

**Figure 3.**
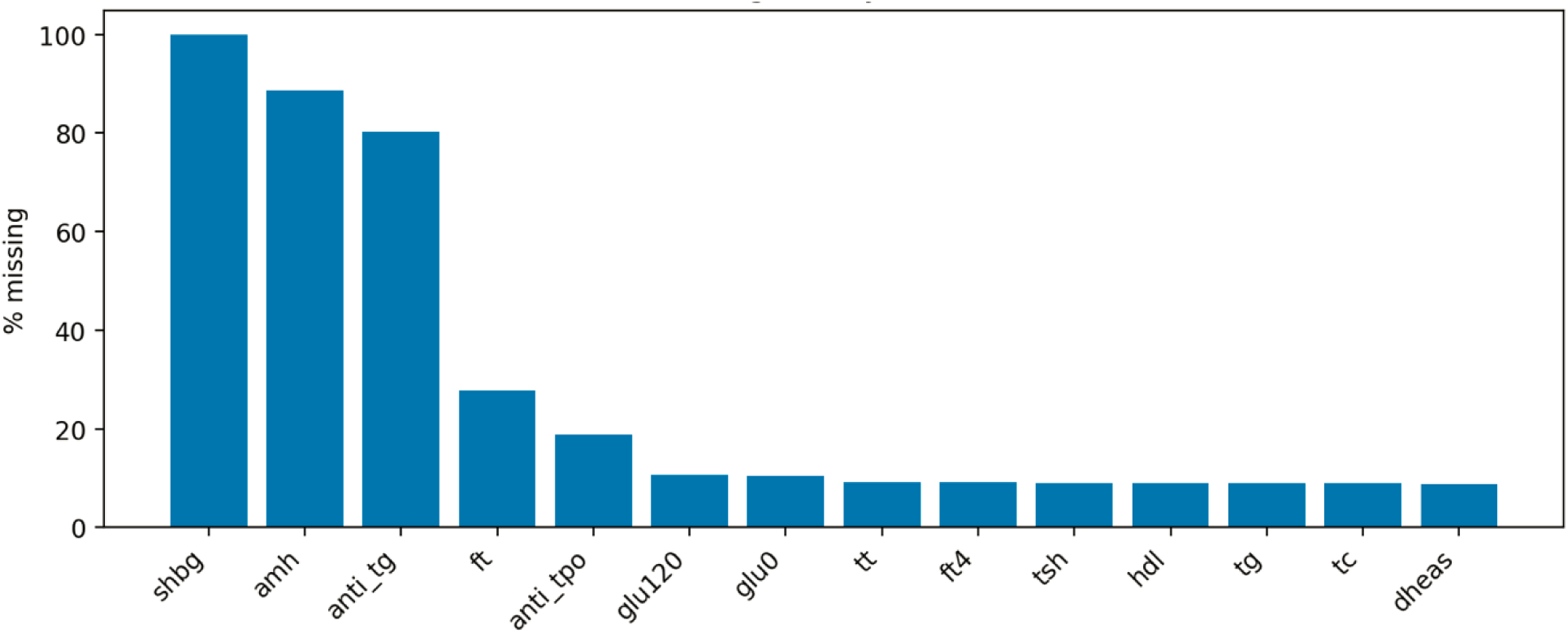
Missingness pattern. SHBG - sex hormone-binding globulin, amh - Anti-Müllerian hormone, anti_tg - anti-thyroglobulin antibodies, ft - free testosterone, anti_tpo - anti-thyroid peroxidase antibodies, glu120 - glucose concentration 120 minutes after oral glucose load, glu0 - fasting glucose, tt - total testosterone, fsh - follicle-stimulating hormone, ft4 - free thyroxine, tsh - thyroid-stimulating hormone, hdl - high-density lipoprotein cholesterol, tg - triglycerides, tc - total cholesterol, dheas - dehydroepiandrosterone sulfate.

Importantly, missingness patterns for key cardiometabolic variables were comparable between TAI-positive and TAI-negative groups, suggesting no major differential missingness that could bias exposure–outcome comparisons.

### 5.2 Primary endpoint analysis

The association between thyroid autoimmunity and the primary cardiometabolic endpoint was evaluated using age-adjusted Firth logistic regression in the complete-case analytic cohort.

Among 1,053 participants with available exposure and outcome data, the primary endpoint (TG/HDL-C ratio >3.5) was observed in 70 women. The event occurred in 4 of 84 TAI-positive participants and 66 of 969 TAI-negative participants.

In the age-adjusted model, thyroid autoimmunity defined by TAI_A was not associated with the primary endpoint. The estimated odds ratio was 0.77 (95% CI 0.21–1.67), indicating no statistically significant association between TAI status and elevated TG/HDL-C ratio.

Adjustment for age did not materially change the direction or magnitude of the effect estimate. The point estimate was below unity, but the confidence interval was wide and crossed 1.0, indicating substantial uncertainty and no evidence of a clinically meaningful increase in cardiometabolic risk associated with thyroid autoimmunity.

A forest plot summarizing the primary regression model is presented in Figure 4.

**Figure 4.**
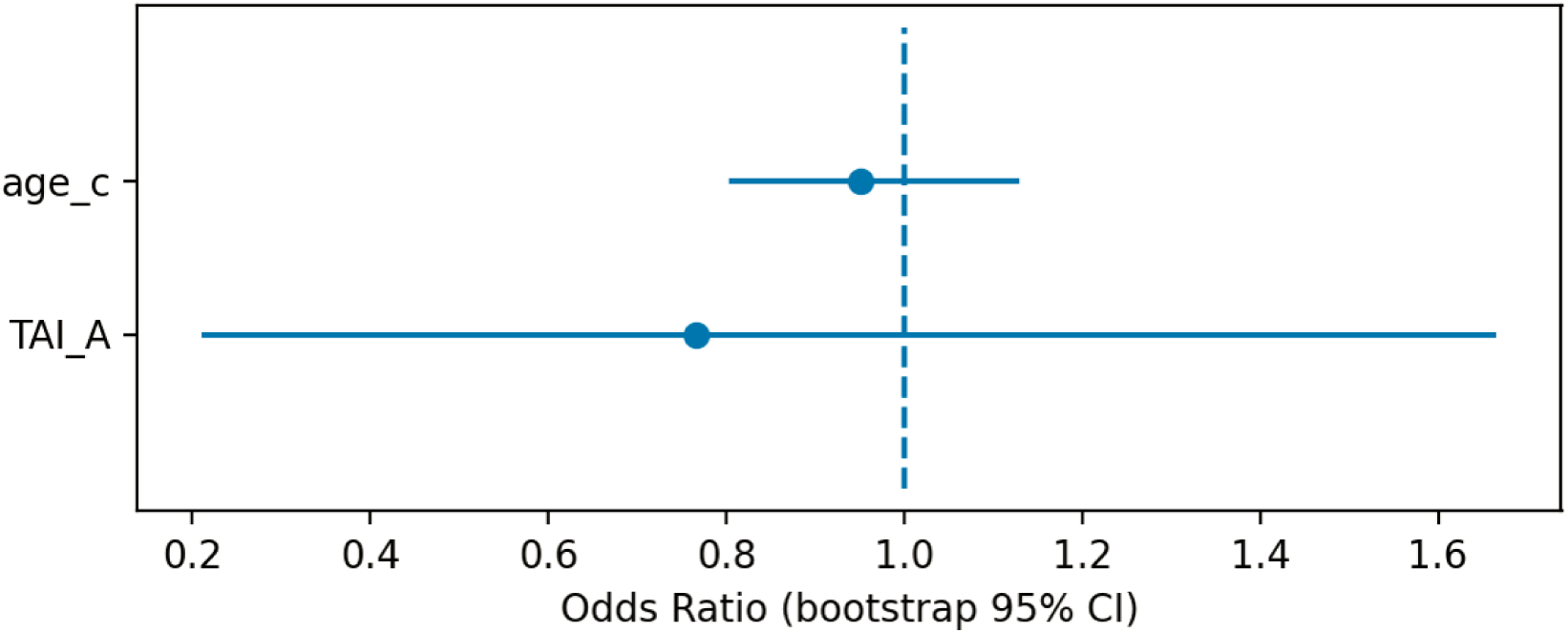
Forest plot of the age-adjusted Firth logistic regression model for the primary endpoint (TG/HDL-C ratio >3.5).

Model calibration analysis demonstrated reasonable agreement between predicted and observed event probabilities, with no evidence of systematic miscalibration (Figure 5).

**Figure 5.**
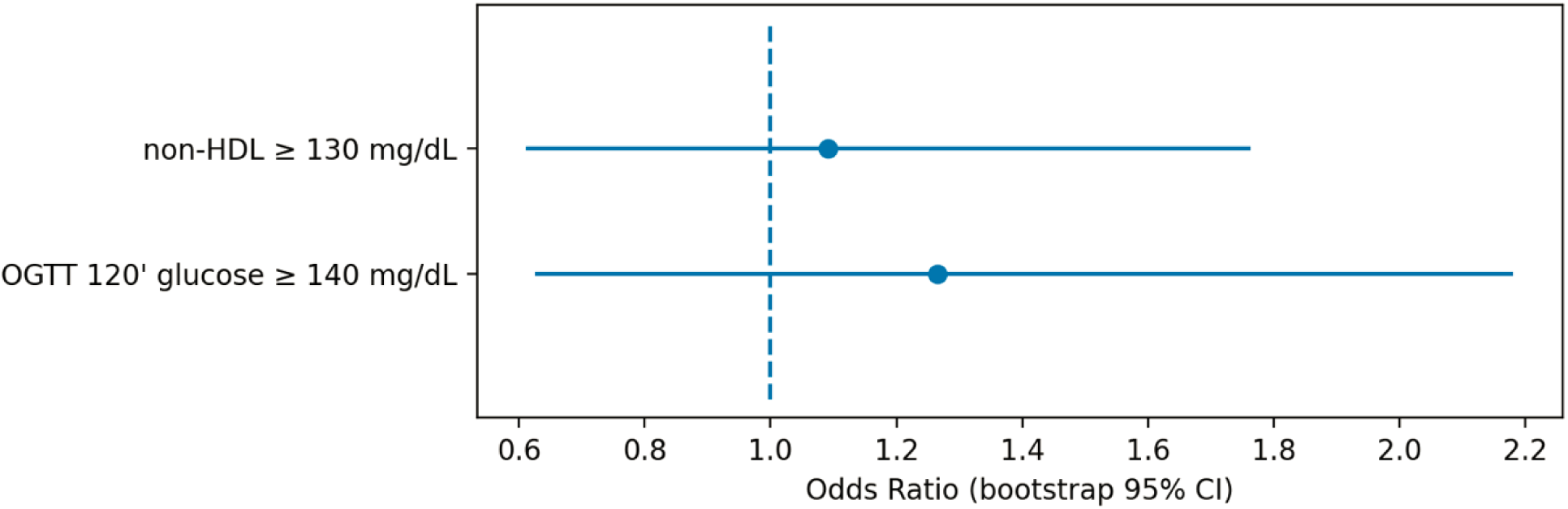
Calibration plot comparing predicted and observed probabilities for the primary endpoint (TG/HDL-C ratio >3.5).

### 5.3 Secondary endpoints

Associations between thyroid autoimmunity and the pre-specified secondary cardiometabolic endpoints were evaluated using age-adjusted Firth logistic regression models.

For elevated non–HDL-C (≥130 mg/dL), the endpoint occurred in 19 of 84 TAI-positive participants and in 205 of 969 TAI-negative participants. In the age-adjusted model, thyroid autoimmunity defined by TAI_A was not significantly associated with elevated non–HDL-C (OR 1.09, 95% CI 0.61–1.76; n = 1053, events = 224).

For impaired glucose tolerance defined as 120-minute OGTT glucose ≥140 mg/dL, the endpoint was observed in 14 of 79 TAI-positive participants and in 142 of 956 TAI-negative participants. Age-adjusted regression analysis similarly did not demonstrate a significant association between TAI_A and impaired glucose tolerance (OR 1.27, 95% CI 0.63–2.18; n = 1035, events = 156).

Effect estimates for both secondary endpoints are summarized in Figure 5. Across both models, point estimates were close to the null value and the confidence intervals included unity, indicating no statistically significant associations between thyroid autoimmunity and these cardiometabolic outcomes.

### 5.4 Androgenic and hormonal group differences

Exploratory comparisons of continuous hormonal and metabolic parameters between TAI-positive and TAI-negative participants were conducted using nonparametric group comparisons with adjustment for multiple testing using the Benjamini–Hochberg false discovery rate (FDR).

A total of 16 candidate markers were evaluated in participants with defined thyroid autoimmunity status (TAI-positive n = 84; TAI-negative n = 971). Among these variables, thyroid-stimulating hormone (TSH) showed the largest standardized group difference and remained statistically significant after FDR correction (Hedges g = 0.90, FDR-adjusted p < 0.001), consistent with the expected alteration of thyroid axis parameters in individuals with thyroid autoimmunity.

In contrast, no androgen-related parameters—including total testosterone, free testosterone, DHEAS, or androstenedione—demonstrated statistically significant differences after FDR adjustment. Similarly, no lipid or glucose metabolism markers showed significant group differences.

These results indicate that, aside from the expected elevation of TSH in TAI-positive individuals, no consistent androgenic or cardiometabolic signal was detected across the evaluated markers. The distribution of effect sizes and adjusted significance levels is illustrated in the volcano plot (Figure 6) and the ranked effect size plot (Figure 7).

**Figure 6.**
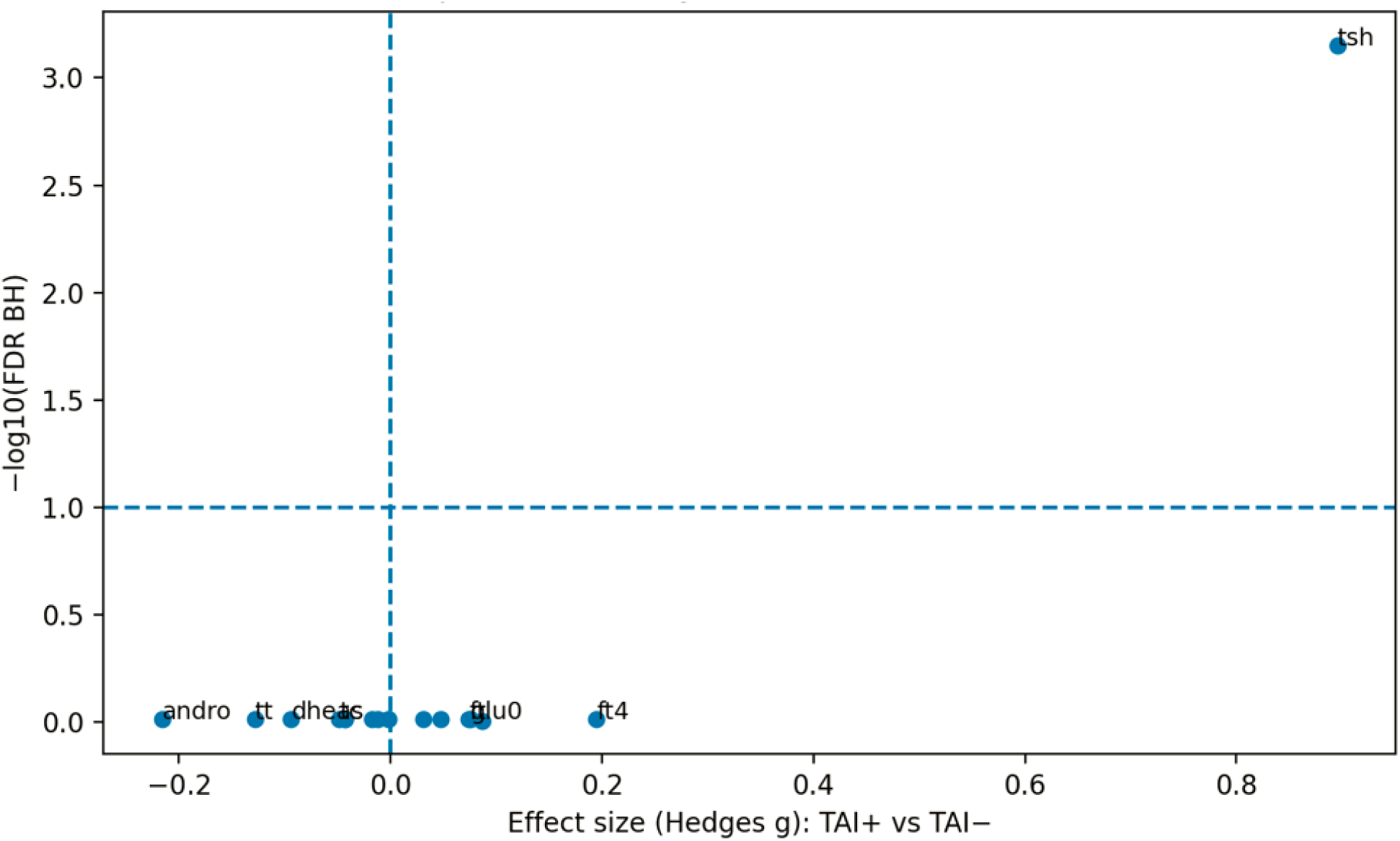
Volcano plot of hormonal and metabolic group differences between TAI-positive and TAI-negative women with PCOS.

**Figure 7.**
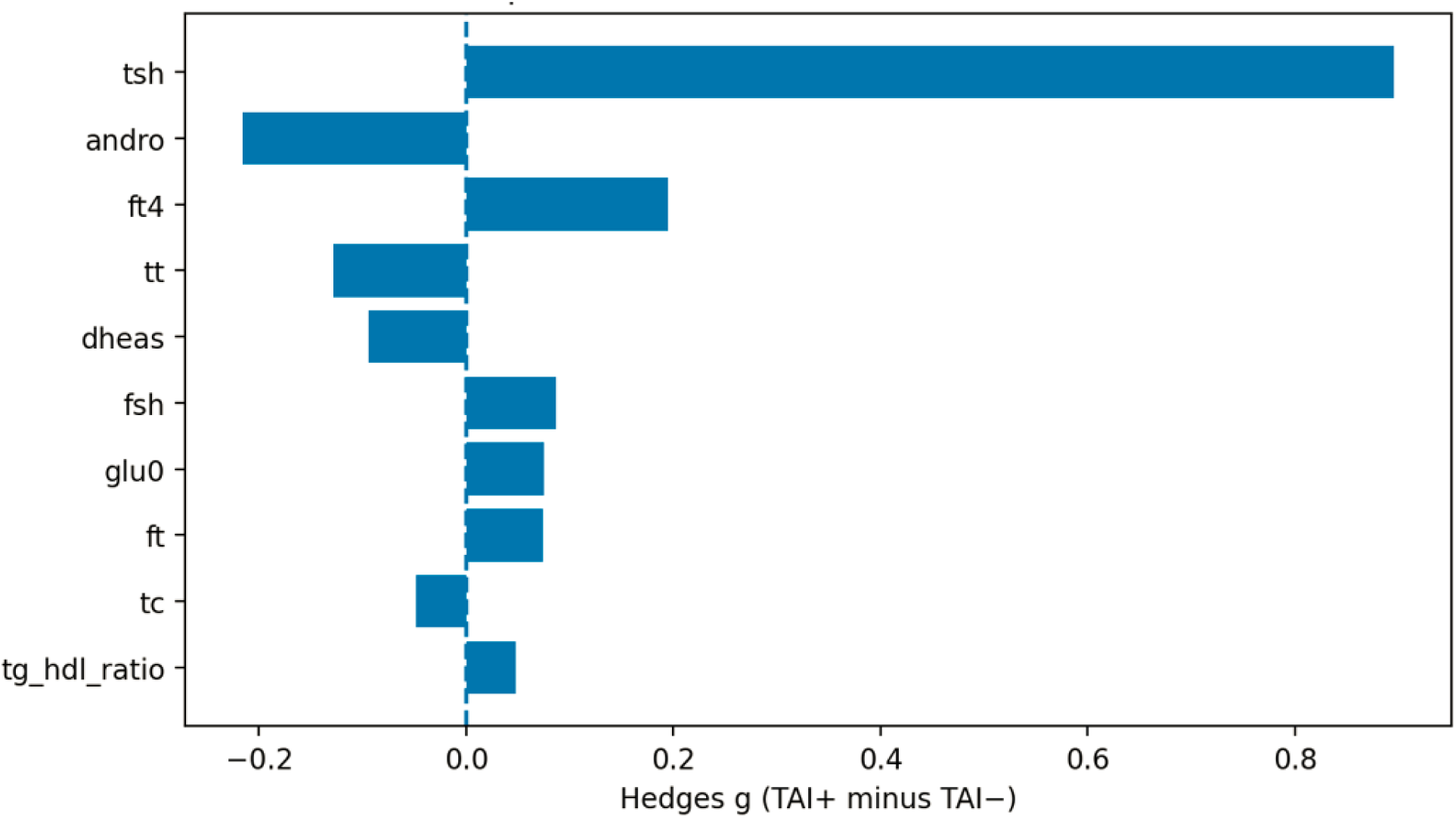
Ranked standardized effect sizes for hormonal and metabolic markers according to thyroid autoimmunity status. tsh - thyroid-stimulating hormone, andro - androstenedione, ft4 - free thyroxine, tt - total testosterone, dheas - dehydroepiandrosterone sulfate, fsh - follicle-stimulating hormone, glu0 - fasting glucose, ft - free testosterone, tc - total cholesterol, tg_hdl_ratio - triglycerides-to-high density lipoprotein cholesterol ratio.

### 5.5 Sensitivity analyses

The robustness of the primary findings was evaluated using several pre-specified sensitivity analyses, including restriction to euthyroid participants, alternative definitions of thyroid autoimmunity, and trimming of extreme values.

When restricting the analysis to euthyroid participants, the association between TAI_A and the primary endpoint remained non-significant (OR 0.50, 95% CI 0.09 - 1.28; n = 1005). Similarly, alternative exposure definitions based on anti-TPO positivity combined with elevated TSH (TAI_B) and high-titer anti-TPO positivity (TAI_C) did not yield statistically significant associations with the primary endpoint (TAI_B: OR 2.87, 95% CI 0.47 - 8.77; TAI_C: OR 1.28, 95% CI 0.34 - 2.90).

Trimming extreme values at the 1st and 99th percentiles also did not materially alter effect estimates for the primary model (OR 0.49, 95% CI 0.08 - 1.20; n = 1033).

Across these analytic scenarios, confidence intervals remained wide and consistently included the null value. Effect estimates across all sensitivity models are summarized in the robustness forest plot (Figure 8).

**Figure 8.**
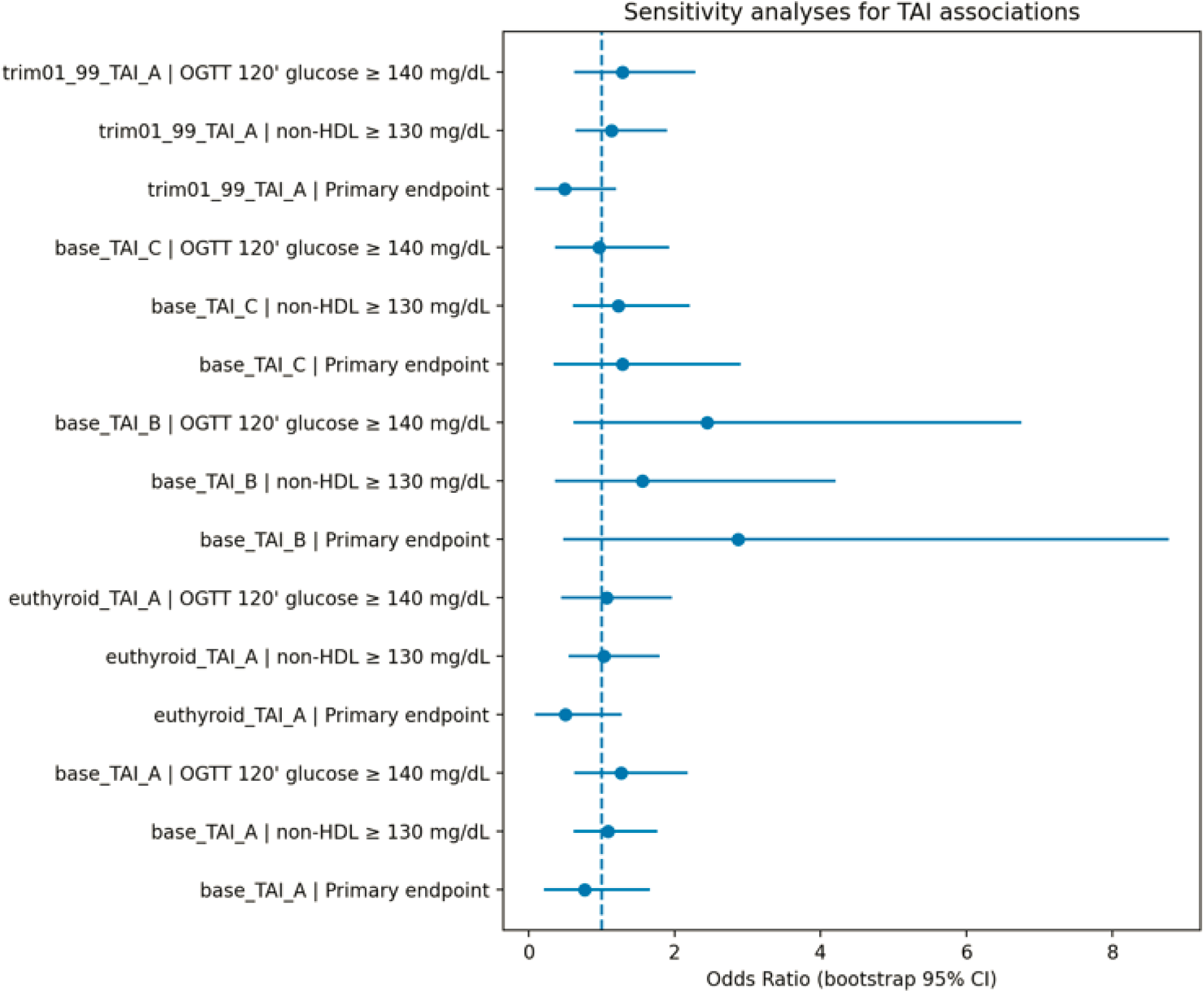
Robustness analysis of the association between thyroid autoimmunity and the primary cardiometabolic endpoint across sensitivity models.

### 5.6 Model diagnostics

Model diagnostics supported appropriate specification of the regression models.

Because the primary analyses were performed using Firth logistic regression, selected diagnostic procedures not directly available for Firth models were evaluated using auxiliary standard logistic models fitted to the same complete-case cohorts.

Calibration plots indicated adequate agreement between predicted and observed risks for the primary and secondary endpoints. Hosmer–Lemeshow goodness-of-fit tests did not indicate evidence of poor calibration in these auxiliary models.

Multicollinearity among predictors was minimal, with variance inflation factors remaining low. Evaluation of influential observations using Cook’s distance did not identify individual observations exerting disproportionate influence on model estimates.

Sensitivity analyses using spline-based modeling of the age covariate produced results consistent with the primary models assuming linear age effects. Detailed diagnostic results are provided in Supplementary Tables S1–S2 and Supplementary Figures S1–S2.

## 6 Discussion

### 6.1 Principal findings

In this pre-specified cross-sectional analysis of a large cohort of women with PCOS, thyroid autoimmunity—defined using several a priori criteria—was not associated with a more adverse cardiometabolic or androgenic phenotype.

No significant association was observed between thyroid autoimmunity and the primary endpoint (TG/HDL-C >3.5), nor with secondary endpoints including non-HDL-C ≥130 mg/dL and 120-minute OGTT glucose ≥140 mg/dL. Effect estimates were modest, confidence intervals included the null value, and findings remained consistent across multiple sensitivity analyses, including alternative definitions of thyroid autoimmunity, restriction to euthyroid participants, and trimming of extreme values.

Exploratory comparisons of continuous hormonal and metabolic parameters, performed with false discovery rate adjustment, did not identify meaningful differences between TAI-positive and TAI-negative women, apart from expected alterations in thyroid axis parameters, most notably higher TSH levels in individuals with thyroid autoimmunity.

Taken together, these findings do not provide evidence that thyroid autoimmunity delineates a metabolically or androgenically more severe PCOS phenotype in this cohort.

### 6.2 Interpretation in the context of previous literature

Thyroid autoimmunity has been frequently reported in women with PCOS, and several previous studies have suggested a potential association between thyroid autoimmunity and adverse metabolic characteristics, including dyslipidemia, insulin resistance, or impaired glucose tolerance (7,11,12). However, the available evidence remains heterogeneous and in some cases inconsistent, partly reflecting differences in study design, sample size, definitions of thyroid autoimmunity, and adjustment for potential confounders.

For example, in a study by Gawron et al. (13), no significant association was observed between PCOS phenotypes and either subclinical hypothyroidism or the presence of circulating anti-thyroid antibodies. Similarly, in a cohort study of women with PCOS conducted by Skrzyńska et al. (14), thyroid abnormalities were not associated with PCOS phenotypes or with key hormonal parameters.

The present study extends this literature by applying pre-specified exposure definitions of thyroid autoimmunity, a clearly defined primary cardiometabolic endpoint, and a range of sensitivity analyses across multiple analytic scenarios. Despite these methodological precautions, no consistent association between thyroid autoimmunity and cardiometabolic or androgenic features of PCOS was observed.

Several factors may help explain discrepancies with some earlier reports. First, previous studies frequently included participants with mixed thyroid functional states or did not explicitly examine euthyroid subgroups. In the current analysis, restriction to euthyroid participants did not materially alter the results, suggesting that subtle variations in thyroid function are unlikely to account for the absence of association observed here.

Second, heterogeneity in the operational definition of thyroid autoimmunity may contribute to inconsistent findings across studies. By evaluating three definitions—including a high-titer anti-TPO threshold—the present analysis attempted to capture both broader and more stringent characterizations of thyroid autoimmunity. None of these definitions demonstrated a stable association with cardiometabolic endpoints.

Third, some previously reported associations may reflect limited statistical power or residual confounding in smaller cohorts. The present analysis, conducted in a relatively large cohort and incorporating bootstrap-based confidence intervals as well as correction for multiple comparisons, reduces the likelihood that the observed null findings are attributable solely to model misspecification or statistical instability.

Taken together, these results suggest that while thyroid autoimmunity is biologically relevant with respect to thyroid axis alterations, it does not appear to independently delineate a metabolically or androgenically more severe PCOS phenotype in this cohort of women with PCOS.

### 6.3 Clinical implications

From a clinical perspective, these findings may help clarify the interpretation of thyroid autoimmunity in women with PCOS. Because PCOS and thyroid autoimmunity frequently co-occur, clinicians may hypothesize that TAI-positive patients represent a subgroup at higher cardiometabolic or androgenic risk. However, in the present cross-sectional analysis, thyroid autoimmunity was not associated with increased odds of an adverse TG/HDL-C ratio, elevated non-HDL-C, or impaired glucose tolerance.

These results suggest that the presence of anti-TPO antibodies alone should not necessarily be interpreted as a marker of increased metabolic severity in young women with PCOS. While screening for thyroid dysfunction remains clinically appropriate, the present findings do not indicate that thyroid autoimmunity per se identifies a cardiometabolically high-risk PCOS subgroup within this population.

At the same time, these conclusions should be interpreted cautiously. The present study was cross-sectional and focused on relatively young women with PCOS, and therefore cannot address potential long-term metabolic consequences associated with thyroid autoimmunity. Further prospective studies will be required to determine whether thyroid autoimmunity influences cardiometabolic trajectories or clinical outcomes later in life.

### 6.4 Strengths

This study has several methodological strengths.

First, the exposure definitions and cardiometabolic endpoints were specified a priori, which reduces the risk of post hoc hypothesis generation and improves interpretability of the findings.

Second, thyroid autoimmunity was evaluated using multiple pre-specified operational definitions, including a stringent high-titer anti-TPO threshold. This approach allowed the robustness of the results to be assessed across alternative definitions of thyroid autoimmunity.

Third, the analysis incorporated several sensitivity strategies, including restriction to euthyroid participants, trimming of extreme values, and bootstrap-based confidence interval estimation. Across these analytic scenarios, the results remained consistent and did not indicate a stable association between thyroid autoimmunity and cardiometabolic endpoints.

Fourth, exploratory hormonal and metabolic comparisons were evaluated using correction for multiple testing with the Benjamini–Hochberg false discovery rate procedure, reducing the risk of false-positive findings.

Finally, the analytical workflow was fully reproducible and implemented using version-controlled scripts with clearly documented preprocessing rules, model diagnostics, and computational environment, which enhances transparency and reproducibility of the study.

### 6.5 Limitations

Several limitations of this study should be acknowledged.

First, the cross-sectional design precludes causal inference and does not allow assessment of longitudinal cardiometabolic outcomes. The present analysis therefore cannot determine whether thyroid autoimmunity influences long-term metabolic trajectories in women with PCOS.

Second, body mass index (BMI) was not available in the dataset and could not be included in the regression models. Given the central role of adiposity in both PCOS pathophysiology and cardiometabolic risk, residual confounding related to obesity cannot be excluded.

Third, information on thyroid hormone replacement therapy, including levothyroxine treatment, was not available in the dataset. As a result, sensitivity analyses excluding participants receiving thyroid hormone therapy could not be performed. Because individuals treated with levothyroxine may differ metabolically from untreated individuals with thyroid autoimmunity, the absence of treatment data represents an additional source of potential confounding.

Fourth, some androgen-related parameters were not available with sufficient completeness to construct composite indices such as the free androgen index (e.g., due to missing SHBG measurements). This may limit the granularity of androgenic phenotype characterization in the present cohort.

Fifth, the number of events for the primary endpoint was relatively small in the TAI-positive group, which contributes to wide confidence intervals and limited precision of effect estimates despite the use of Firth logistic regression.

Finally, the study population consisted of women evaluated in a single clinical setting and primarily represented relatively young patients with PCOS. These characteristics may limit the generalizability of the findings to other populations or to older women with PCOS.

## 7 Conclusion

In this pre-specified cross-sectional analysis of women with PCOS, thyroid autoimmunity was not associated with an adverse cardiometabolic or androgenic phenotype. Across multiple operational definitions of thyroid autoimmunity and several sensitivity analyses, no consistent associations were observed with elevated TG/HDL-C ratio, non-HDL-C, impaired glucose tolerance, or androgen-related parameters.

Although thyroid autoimmunity was associated with expected alterations in thyroid axis markers, it did not delineate a subgroup of women with PCOS characterized by greater metabolic or androgenic severity under the studied conditions. These findings suggest that anti-TPO positivity alone should not necessarily be interpreted as an indicator of increased cardiometabolic risk in young women with PCOS.

Further prospective studies incorporating longitudinal follow-up and comprehensive metabolic profiling are needed to clarify whether thyroid autoimmunity influences long-term cardiometabolic outcomes in this population.

## Data Availability

The data that support the findings of this study are not publicly available due to privacy regulations and institutional data protection policies. However, anonymized data may be made available from the corresponding author upon reasonable request and with permission of the Wroclaw Medical University. The code used for data analysis is publicly available at: https://github.com/npiorkowska-science/pcos-thyroid-autoimmunity-analysis

## Acknowledgments

The authors acknowledge Wroclaw Medical University for providing access to the clinical data used in this study. The authors also thank collaborators who contributed to the development of the methodological framework and provided valuable feedback during the preparation of the manuscript.

## Author Contributions

Natalia Piórkowska (NP): Conceptualization, Methodology, Software, Formal analysis, Data curation, Investigation, Writing – original draft, Writing – review & editing.

Lech Madeyski (LM): Methodology, Software, Validation, Supervision, Writing – original draft, Writing – review & editing.

Marcin Leśniewski (ML): Preparation of the clinical database. Grzegorz Franik (GF): Patient diagnosis, Clinical evaluation.

Anna Bizoń (AB): Literature review, Writing – review & editing, Project administration.

All authors approved the final version of the manuscript and agreed to be accountable for all aspects of the work.

## Ethics Statement

The study was conducted in accordance with the principles of the Declaration of Helsinki and Good Clinical Practice guidelines. Ethical approval for the research project entitled “Molecular studies useful for the assessment of metabolic disorders occurring in patients with polycystic ovary syndrome” was granted by the Bioethics Committee at Wroclaw Medical University (Opinion No. KB-254/2021, issued on 31 March 2021).

The project was conducted under the supervision of dr hab. Anna Bizoń at Wroclaw Medical University. The analysis used anonymized clinical data, and all procedures were performed in accordance with institutional regulations regarding the protection of personal data.

## Data and Code Availability

The code used for data preprocessing, feature engineering, model development, and statistical analysis is publicly available in a GitHub repository: https://github.com/npiorkowska-science/pcos-thyroid-autoimmunity-analysis

The repository contains scripts required to reproduce the analytical pipeline, including data preprocessing procedures, model implementation, and evaluation workflows.

Due to privacy regulations and institutional data protection policies, the original clinical dataset cannot be publicly shared. However, the repository includes documentation describing the structure of the dataset and the variables required to reproduce the analysis using compatible data sources.

## Figures

Figure 1. Distribution of thyroid and cardiometabolic

Figure 2. Androgen distributions

Figure 3. Missingness pattern

Figure 4. Forest plot primary model

Figure 5. Forest plot secondary endpoints

Figure 6. Volcano plot of hormonal and metabolic group differences between TAI-positive and TAI-negative women with PCOS.

Figure 7. Ranked standardized effect sizes for hormonal and metabolic markers according to thyroid autoimmunity status.

Figure 8. Robustness analysis of the association between thyroid autoimmunity and the primary cardiometabolic endpoint across sensitivity models.

## Tables

Table S1. Baseline characteristics

## SUPPLEMENTARY MATERIALS

### Supplementary Figures

Supplementary Figure S1. Calibration plot for the primary regression model.

Supplementary Figure S2. Influence diagnostics for the primary regression model.

### Supplementary Tables

Supplementary Table S1. Hosmer–Lemeshow goodness-of-fit tests for auxiliary logistic regression models used in model diagnostics.

Supplementary Table S2. Variance inflation factors (VIF) for predictors included in regression models.

## References

1. Azziz R. Polycystic Ovary Syndrome. Obstet. Gynecol. 2018;132(2):321–336.

2. Goodarzi MO, Dumesic DA, Chazenbalk G, Azziz R. Polycystic ovary syndrome: etiology, pathogenesis and diagnosis. Nat. Rev. Endocrinol. 2011;7(4):219–231.

3. Dumesic DA, Oberfield SE, Stener-Victorin E, Marshall JC, Laven JS, Legro RS. Scientific Statement on the Diagnostic Criteria, Epidemiology, Pathophysiology, and Molecular Genetics of Polycystic Ovary Syndrome. Endocr. Rev. 2015;36(5):487–525.

4. Dapas M, Lin FTJ, Nadkarni GN, Sisk R, Legro RS, Urbanek M, Hayes MG, Dunaif A. Distinct subtypes of polycystic ovary syndrome with novel genetic associations: An unsupervised, phenotypic clustering analysis. PLoS Med. 2020;17(6):e1003132.

5. Garelli S, Masiero S, Plebani M, Chen S, Furmaniak J, Armanini D, Betterle C. High prevalence of chronic thyroiditis in patients with polycystic ovary syndrome. Eur. J. Obstet. Gynecol. Reprod. Biol. 2013;169(2):248–251.

6. Romitti M, Fabris VC, Ziegelmann PK, Maia AL, Spritzer PM. Association between PCOS and autoimmune thyroid disease: a systematic review and meta-analysis. Endocr. Connect. 2018;7(11):1158–1167.

7. Suchta K, Zeber-Lubecka N, Grymowicz M, Smolarczyk R, Kulecka M, Hennig EE. Autoimmune Processes and Chronic Inflammation as Independent Risk Factors for Metabolic Complications in Women with Polycystic Ovary Syndrome. Metabolites 2025;15(3):141.

8. Wang J, Yin T, Liu S. Dysregulation of immune response in PCOS organ system. Front. Immunol. 2023;14:1169232.

9. Kwiatkowski J, Akpang N, Ziemkiewicz Z, Zaborowska L, Ludwin A. Molecular Insights into Elevated Autoantibodies in Polycystic Ovary Syndrome: Mechanisms and Clinical Implications. Int. J. Mol. Sci. 2025;26(17):8192.

10. Hu X, Chen Y, Shen Y, Zhou S, Fei W, Yang Y, Que H. Correlation between Hashimoto’s thyroiditis and polycystic ovary syndrome: A systematic review and meta-analysis. Front. Endocrinol. 2022;13:1025267.

11. Janssen OE, Mehlmauer N, Hahn S, Offner AH, Gärtner R. High prevalence of autoimmune thyroiditis in patients with polycystic ovary syndrome. Eur. J. Endocrinol. 2004;150(3):363–369.

12. Palomba S, Colombo C, Busnelli A, Caserta D, Vitale G. Polycystic ovary syndrome and thyroid disorder: a comprehensive narrative review of the literature. Front. Endocrinol. 2023;14:1251866.

13. Gawron IM, Baran R, Derbisz K, Jach R. Association of Subclinical Hypothyroidism with Present and Absent Anti-Thyroid Antibodies with PCOS Phenotypes and Metabolic Profile. J. Clin. Med. 2022;11(6):1547.

14. Skrzyńska KJ, Zachurzok A, Gawlik AM. Metabolic and Hormonal Profile of Adolescent Girls With Polycystic Ovary Syndrome With Concomitant Autoimmune Thyroiditis. Front. Endocrinol. 2021;12:708910.

